# Distinct patterns of blood cytokines beyond a cytokine storm predict mortality in COVID-19

**DOI:** 10.1101/2021.05.04.21256497

**Authors:** Christian Herr, Sebastian Mang, Bahareh Mozafari, Katharina Günther, Thimoteus Speer, Martina Seibert, Sanjay Kumar Srikakulam, Christoph Beisswenger, Felix Ritzmann, Andreas Keller, Rolf Müller, Sigrun Smola, Dominic Eisinger, Michael Zemlin, Guy Danziger, Thomas Volk, Sabrina Hörsch, Marcin Krawczyk, Frank Lammert, Thomas Adams, Gudrun Wagenpfeil, Michael Kindermann, Constantin Marcu, Zuhair Wolf Dietrich Ataya, Marc Mittag, Konrad Schwarzkopf, Florian Custodis, Daniel Grandt, Harald Schäfer, Kai Eltges, Philipp M. Lepper, Robert Bals, CORSAAR study group

**Affiliations:** Department of Internal Medicine V – Pulmonology, Allergology and Critical Care Medicine, Saarland University, D-66421 Homburg, Germany; Department of Internal Medicine IV, Nephrology and Hypertension & Translational Cardio-Renal Medicine, Saarland University, Homburg/Saar, Germany Saarland; Clinical Bioinformatics, Saarland University, 66421 Homburg, Germany; Helmholtz-Institut für Pharmazeutische Forschung Saarland, 66123 Saarbrücken, Germany; Institute for Virology, Saarland University, 66421 Homburg, Germany; Myriad RBM Inc. 3300 Duval Rd. Austin, Texas 78759, USA; Department for General Pediatrics and Neonatology, Saarland University, 66421 Homburg, Germany; Clinic for Anesthesiology, Intensive Care Medicine and Pain Therapy, Saarland University, D-66421 Homburg, Germany; Department of Internal Medicine II, Saarland University, D-66421 Homburg, Germany; Institute for Medical Biometry, Epidemiology and Medical Informatics, Saarland University, Campus Homburg/Saar, Germany; Caritas Klinikum Saarbrücken St. Theresia, Saarbrücken, Saarland, Germany; Innere Medizin und Internistische Intensivmedizin, Caritas Klinikum, Saarbrücken St. Josef, Dudweiler, Saarland, Germany; Klinikum Saarbrücken, 66119 Saarbrücken, Germany; Department of Internal Medicine and Pulmonology, SHG-Kliniken Völklingen, Völklingen, Saarland, Germany

**Keywords:** biomarker, inflammation, SARS-CoV2

## Abstract

**Background:** COVID-19 comprises several severity stages ranging from oligosymptomatic disease to multi-organ failure and fatal outcomes. The mechanisms why COVID-19 is a mild disease in some patients and progresses to a severe multi-organ and often fatal disease with respiratory failure are not known. Biomarkers that predict the course of disease are urgently needed. The aim of this study was to evaluate a large spectrum of established laboratory measurements.

**Patients and methods:** Patients from the prospective PULMPOHOM and CORSAAR studies were recruited and comprised 35 patients with COVID-19, 23 with conventional pneumonia, and 28 control patients undergoing elective non-pulmonary surgery. Venous blood was used to measure the serum concentrations of 79 proteins by Luminex multiplex immunoassay technology. Distribution of biomarkers between groups and association with disease severity and outcomes were analyzed.

**Findings:** The biomarker profiles between the three groups differed significantly with elevation of specific proteins specific for the respective conditions. Several biomarkers correlated significantly with disease severity and death. Uniform manifold approximation and projection (UMAP) analysis revealed a significant separation of the three disease groups and separated between survivors and deceased patients. Different models were developed to predict mortality based on the baseline measurements of several protein markers.

**Interpretation:** Several newly identified blood markers were increased in patients with severe COVID-19 (AAT, EN-RAGE, ICAM-1, myoglobin, SAP, TIMP-1, vWF, decorin, HGF, MMP7, PECAM-1) or in patients that died (FRTN, SCF, TIMP-1, CA-9, CEA, decorin, HGF). The use of established assay technologies allows for rapid translation into clinical practice.

**Funding:** No role of the funding source.

## Introduction

Coronavirus disease 2019 (COVID-19) is a novel illness and a rapidly evolving pandemic with large impact on global health. COVID-19 is caused by “severe acute respiratory syndrome coronavirus 2” (SARS-CoV-2), a member of the Coronaviridae family that is regarded as a spill-over of an animal coronavirus to humans [1]. At the time of submission of this work, SARS-CoV-2 has caused 120 million diagnosed infections and 6 million known deaths. The disease course is complex, increasing disease severity is associated with respiratory and extra-pulmonary organ manifestations. Most of the infected individuals have mild flu-like symptoms, nevertheless, about 20% of individuals develop more severe illness due to viral pneumonia, often resulting in the need for hospitalization. Often, the disease progresses, and patients need invasive life support. The mechanisms why COVID-19 is a mild disease in some patients and progresses to a severe multi-organ and often fatal disease with respiratory failure are currently unknown.

The natural history and the pathomechanisms of COVID-19 involve complex mechanisms including infection and viral replication in target cells and a broad dysregulation of host defense and immune mechanisms. Several studies have evaluated blood transcriptional data and highlighted the importance of dysregulation of innate [2] or adaptive immunity including activation of type 1 (t-bet, interferon-γ), type-2 (GATA) or type 3 (RORγt, IL-17) immune pathways [3, 4].

Elevation of routine inflammation markers, such as erythrocyte sedimentation rate, C-reactive protein (CRP) or procalcitonin (PCT), is frequently but not exclusively observed in the critically ill COVID-19 patient [5]. The term “cytokine storm” has been used to describe the elevation of several acute-phase proteins such as CRP, interleukin-6 (IL-6) or ferritin [6, 7]. Individual markers or patterns have been associated with disease severity or lethality [8–10].

There is a clinical need to identify markers that indicate the risk of disease progression or fatal outcome. The aim of the present study was to identify clinically relevant serum biomarkers from a large spectrum of established laboratory measurements. The marker panel applied in the present study consists of mediators of inflammation and immunity, which are not yet part of clinical routine testing but have been shown to be involved in cellular or humoral immune responses. We measured serum levels of a large panel of cytokines, chemokines, acute phase reactants, growth factors, tissue remodeling enzymes and clinical laboratory parameters in blood samples of hospitalized COVID-19 patients and compared them to control individuals undergoing non-thoracic surgery as well as patients with community-acquired pneumonia.

## Methods

### Study population

The present analysis used the baseline data and follow up data of the COVID-19 cohort CORSAAR (n□= 35) and its control cohort PULMOHOM, which are multi-center studies focusing on pathomechanisms and the role of risk factors in COVID-19 and other inflammatory lung diseases. Within the PULMOHOM cohort study, 23 patients with pneumonia and 28 control patients undergoing elective non-pulmonary surgery have been included. Patients were recruited at the Saarland University Hospital, the Caritas Klinikum Saarbrücken, the Klinikum Saarbrücken Winterberg, and the SHG-Kliniken Völklingen. The studies have been approved by the ethics committee of the Ärztekammer des Saarlandes, and all patients or their legal representatives gave their informed consent. Basic and anthropomorphic characteristics, the prevalence of comorbidities, and vital parameters were assessed based on measurements, questionnaires, and standardized interviews.

### Biosamples

Blood samples were drawn after obtaining informed consent from the patient and processed within 4 hours (time-to-centrifugation). Blood was centrifuged for 20 minutes at 2500 g at 4°C in a swing out rotor with break. The serum supernatant was separated immediately after centrifugation under aseptical conditions and transferred into cryo-vials. The vials were integrated into our local biobank (−80°C storage and Biobank Information System (BIMS)) for long term storage. The freezers of the biobank are temperature monitored and access controlled. Upon selection for analysis, the samples were sent on dry ice to the corresponding labs.

### Multiplex immunoassay analysis

All samples were stored at less than −70°C until tested. Samples were thawed at room temperature, vortexed, spun at 3700 x g for 5 min for clarification and transferred to a master microtiter plate. Using automated pipetting, an aliquot of each sample was added to individual microsphere multiplexes of the selected Multi Analyte Profile (MAP) and blocker. This mixture was thoroughly mixed and incubated at room temperature for 1 hour. Multiplexed cocktails of biotinylated reporter antibodies were added robotically and after thorough mixing, incubated for an additional hour at room temperature. Multiplexes were labelled using an excess of streptavidin-phycoerythrin solution, thoroughly mixed and incubated for 1 hour at room temperature. The volume of each multiplexed reaction was reduced by vacuum filtration and washed thrice. After the final wash, the volume was increased by addition of buffer for analysis using a Luminex instrument and the resulting data interpreted using proprietary software developed by Myriad RBM. For each multiplex, both calibrators and controls were included on each microtiter plate. Eight-point calibrators to form a standard curve were run in the first and last column of each plate and controls at three concentration levels were run in duplicate. Standard curve, control, and sample QC were performed to ensure proper assay performance. Study sample values for each of the analytes were determined using 4 and 5 parameter logistics, weighted and non-weighted curve fitting algorithms included in the data analysis package.

The following assays were used: adiponectin, alpha-1-antitrypsin (AAT), alpha-2-macroglobulin (A2Macro), apolipoprotein(a) (Lp(a)), beta-2-microglobulin (B2M), brain-derived neurotrophic factor (BDNF), c-reactive protein (CRP), complement C3 (C3), EN-RAGE, eotaxin-1, factor VII, ferritin (FRTN), fibrinogen, granulocyte-macrophage colony-stimulating factor (GM-CSF), haptoglobin, immunoglobulin a (IgA), immunoglobulin m (IgM), intercellular adhesion molecule 1 (ICAM-1), interferon gamma (IFN-gamma), interleukin-1 alpha (IL-1 alpha), interleukin-1 beta (IL-1 beta), interleukin-1 receptor antagonist (IL-1ra), interleukin-2 (IL-2), interleukin-3 (IL-3), interleukin-4 (IL-4), interleukin-5 (IL-5), interleukin-6 (IL-6), interleukin-7 (IL-7), interleukin-8 (IL-8), interleukin-10 (IL-10), interleukin-12 Subunit p40 (IL-12p40), interleukin-12 subunit p70 (IL-12p70), interleukin-17 (IL-17), interleukin-18 (IL-18), macrophage inflammatory protein-1 alpha (MIP-1 alpha), macrophage inflammatory protein-1 beta (MIP-1 beta), matrix metalloproteinase-3 (MMP-3), matrix metalloproteinase-9 (MMP-9), monocyte chemotactic protein 1 (MCP-1), myoglobin, plasminogen activator inhibitor 1 (PAI-1), pulmonary and activation-regulated chemokine (PARC), serum amyloid p-component (SAP), stem cell factor (SCF), t-cell-specific protein RANTES (RANTES), thyroxine-binding globulin (TBG), tissue inhibitor of metalloproteinases 1 (TIMP-1), tumor necrosis factor alpha (TNF-alpha), tumor necrosis factor beta (TNF-beta), tumor necrosis factor receptor 2 (TNFR2), vascular cell adhesion molecule-1 (VCAM-1), vascular endothelial growth factor (VEGF), vitamin d-binding protein (VDBP), von Willebrand factor (vWF)), AXL receptor tyrosine kinase (AXL), chemokine CC-4 (HCC-4), FASLG receptor (FAS), hepatocyte growth factor (HGF), TNF-related apoptosis-inducing ligand receptor 3 (TRAIL-R3)), alpha-fetoprotein (AFP), cancer antigen 125 (CA-125), cancer antigen 19-9 (CA-19-9), carcinoembryonic antigen (CEA), human chorionic gonadotropin beta (hCG), neuron-specific enolase (NSE)), matrix metalloproteinase-1 (MMP-1), matrix metalloproteinase-7 (MMP-7), matrix metalloproteinase-9, total (MMP-9, total), angiopoietin-1 (ANG-1), carbonic anhydrase 9 (CA-9), decorin, interleukin-18-binding protein (IL-18bp), platelet endothelial cell adhesion molecule (PECAM-1), pulmonary surfactant-associated protein D (SP-D).

### Statistical analyses

Continuous variables are presented as mean ± standard deviation when normally distributed or as median (standard deviation). Qualitative variables are presented as absolute and relative frequencies. Statistical differences between three groups considering continuous or categorical variables have been established using one-way Analysis of Variance (ANOVA, assuming normality) with Bonferroni correction as post-hoc test, Kruskal-Wallis test or chi-squared test, respectively. Differences between two groups were analyzed by two-sided T-test. For bivariate correlations, the Pearson’s R is reported. A correlation matrix was generated by using R language (version 3.6.1) and plotted using ggplot2 library. For plotting, only the absolute correlation values >0.3 were retained. A linear discriminant analysis was performed to test whether the serum markers identified above can be used to build a model that indicates increased risk of mortality. Neural network analysis (SPPS module multilayer perceptron, MLP, one hidden layer) was performed using “death” as dependent variable and the biomarkers with different serum concentrations in survivors vs. non-survivors as covariates. A two-sided *P* value of less than 0.05 was considered statistically significant. Statistical analyses were performed using SPSS version 27.0. Dimension reduction and visualization of high dimensional profiles has additionally been carried out using the Uniform Manifold Approximation and Projection (UMAP) by using the R umap package. Input data were z-scaled before computing the UMAP. The number of components was set to 2, and the Euclidean distance was used. The computations for the UMAP used R 4.0.3 GUI 1.73 Catalina build (7892).

## Results

### The blood inflammatory profile differs between COVID-19 and conventional pneumonia

As a first step we compared the concentrations of blood mediators between patients with pneumonia, COVID-19 and a control group of patients undergoing non-pulmonary surgery. Table 1 summarizes the main characteristics of the patients in these groups and shows that the three groups did not differ with respect to age or body mass index (BMI). Table 1S summarizes the clinical characteristics of all patients. Only male patients were recruited for the control and pneumonia groups to avoid sex-related changes of metabolic parameters. We analyzed the data from the measurements and found that the concentrations of IL-1-alpha, IL-3, IL-4, IL-7, IL-12p70, TNF-beta, and TNF-alpha were below the detection threshold. These markers were excluded from further analysis.

**Table 1:** Distribution of age, sex, and BMI between the study groups. Data are median (SD) and n (%). ** indicates p < 0.01 as calculated by Kruskal-Wallis test.

| Item | Control | Pneumonia | COVID-19 | Stat. |
| --- | --- | --- | --- | --- |
| Patient number | 28 | 23 | 35 | - |
| Sex, % male | 100 | 100 | 74 | ** |
| Age | 62.42 (2.14) | 64.61 (3.11) | 63.86 (3.18) | - |
| BMI | 26.30 (0.99) | 27.27 (2.22) | 27.08 (1.31) | - |

A direct comparison of results from the three groups showed distinct patterns of elevated venous markers when analyzed by ANOVA with Bonferroni correction as post-hoc test. The three groups differ with regard to the concentrations of various cytokines: CRP, AAT, B2M, BDNF, EN-RAGE, fibrinogen, haptoglobin, IgA, IgM, ICAM-1, IL-1b, IL-8, MIP-1alpha, MMP-3, MMP-9, MCP-1, myoglobin, PAI-1, PARC, SAP, SCF, RANTES, TIMP-1, VCAM-1, vWF, AXL, HCC-4, HGF, MMP-7, MMP-9, NSE, PECAM-1, SP-D, A2Macro, IL-18, CA-125, CA-9, decorin, IL-18bp and TRAIL-R3. The data are summarized in Table 2. Figure 1A displays a selection of markers with significant differences.

**Table 2:**
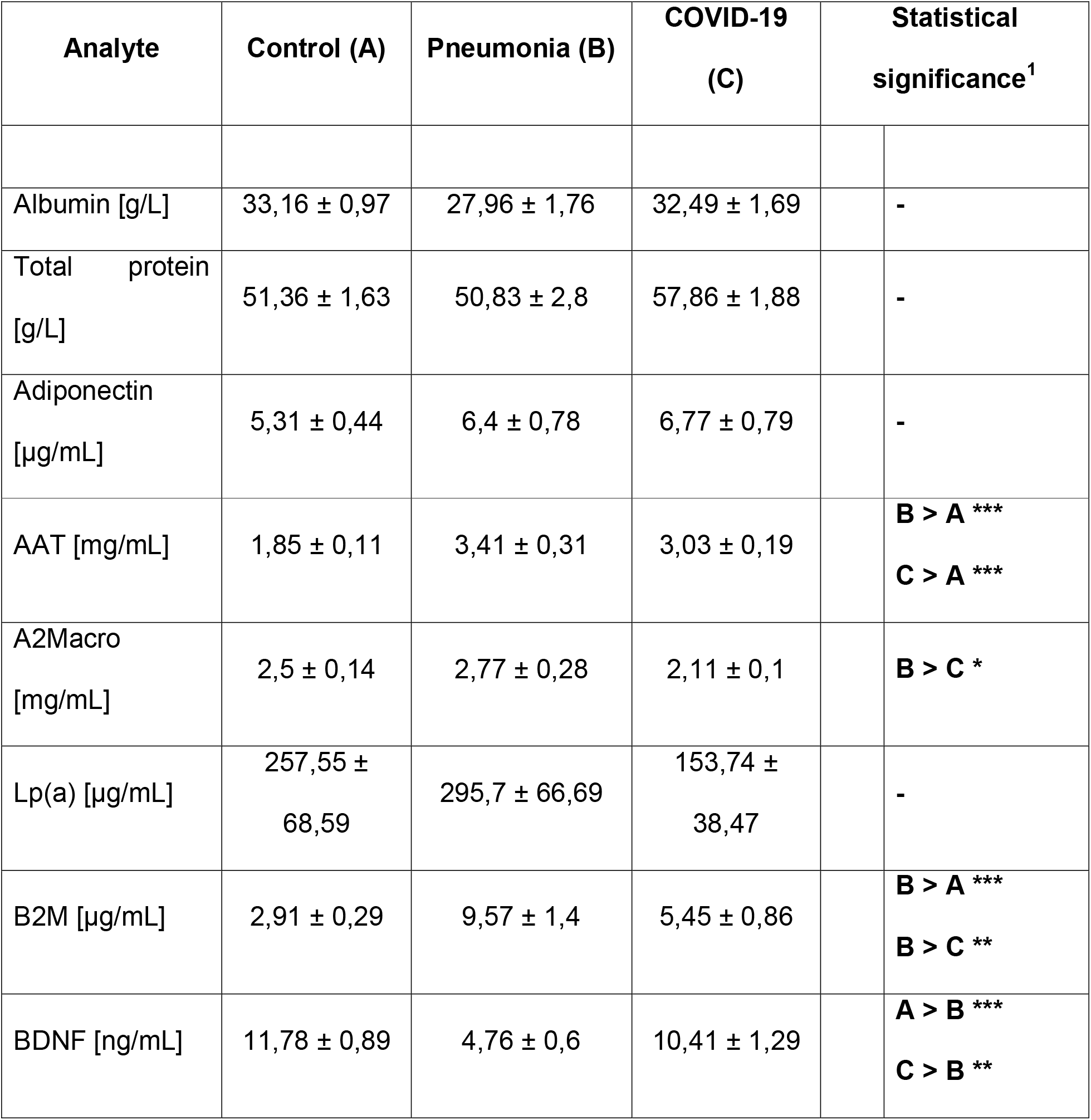
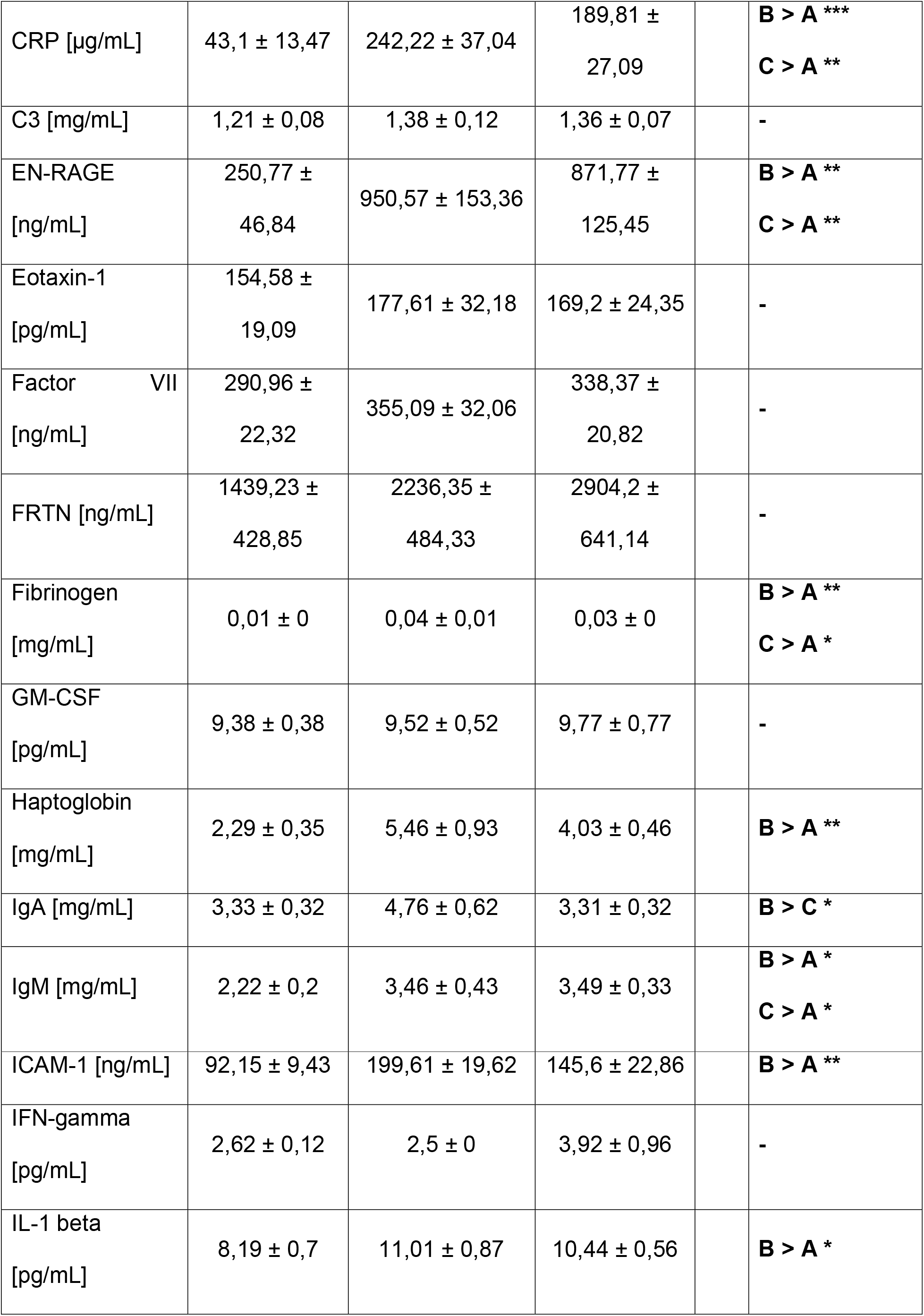
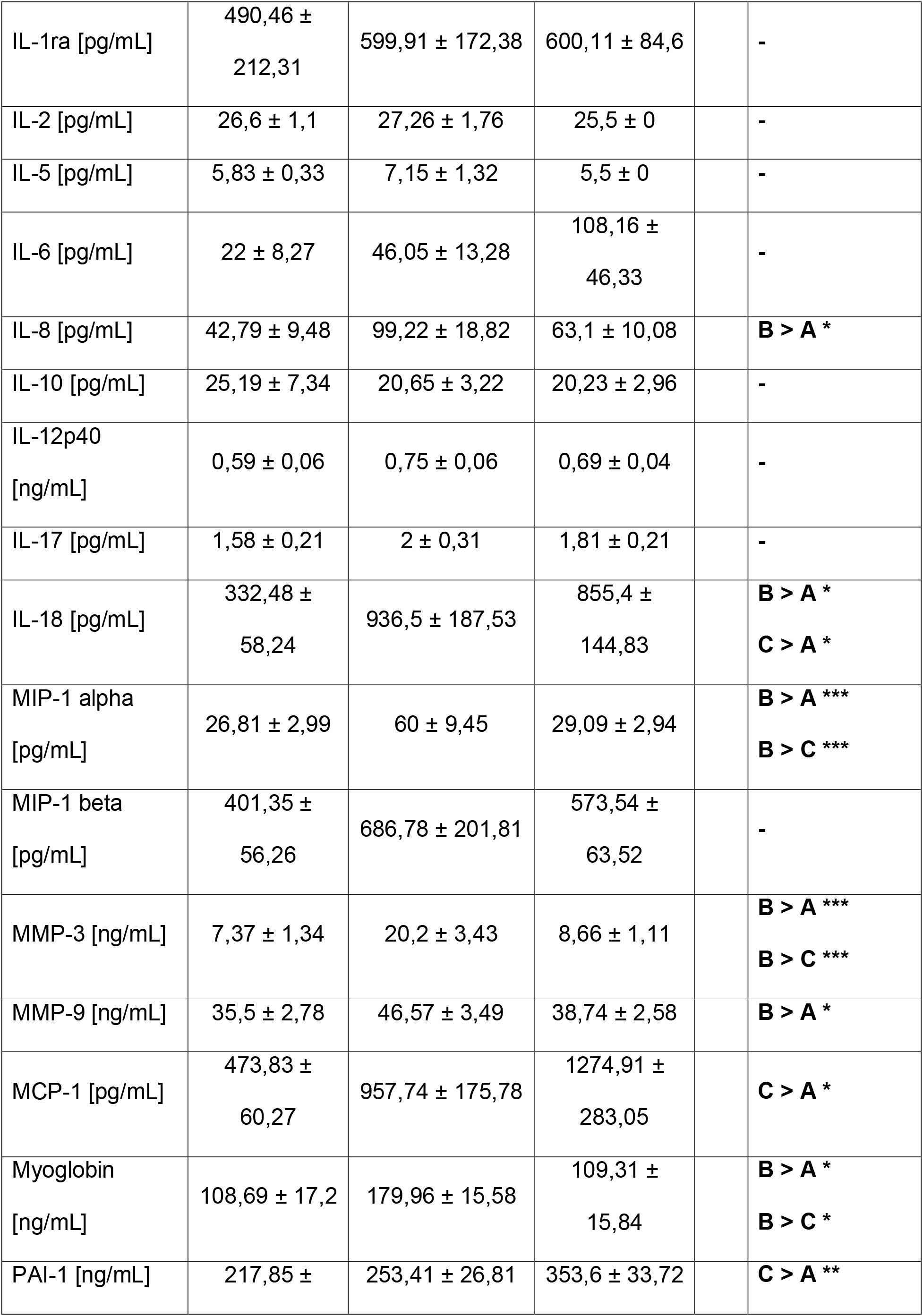
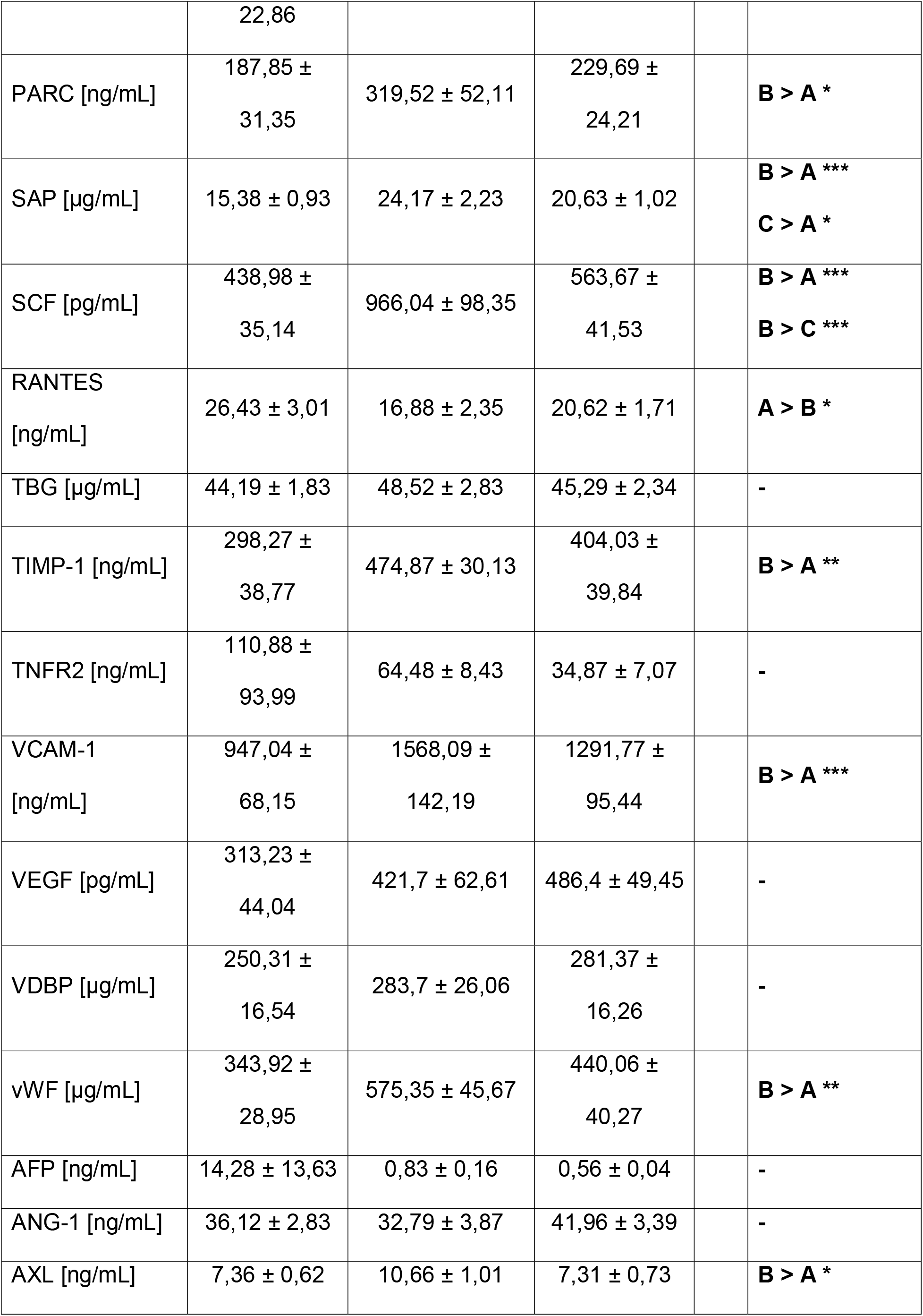
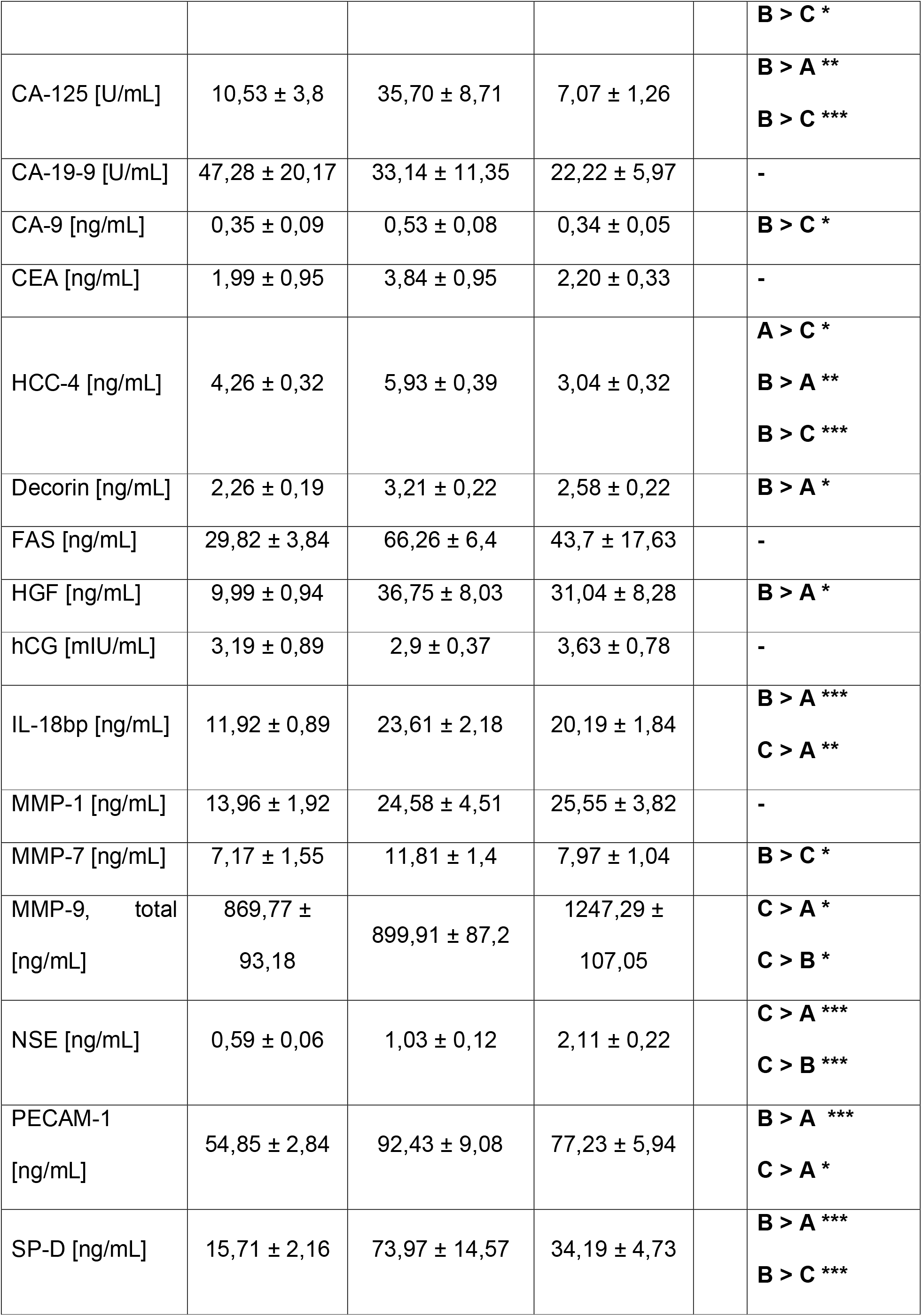
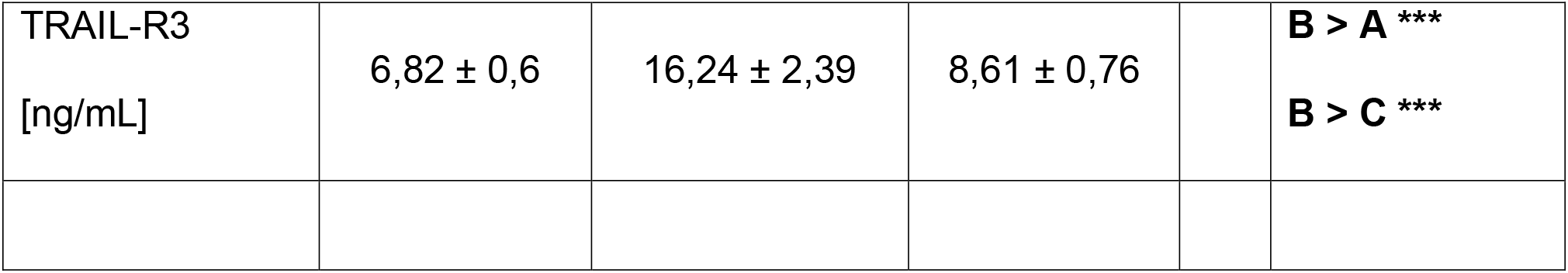
Serum concentrations of blood proteins in patients with pneumonia, COVID-19 and control patients without lung disease undergoing elective surgery. Data are median ± SD. * indicates p < 0.05, ** p < 0.005, *** p < 0.001 as calculated using one-way Analysis of Variance (ANOVA, assuming normality) with Bonferroni correction as post-hoc test. The column on the right indicated differences between groups.

| Analyte | Control (A) | Pneumonia (B) | COVID-19 (C) | Statistical significance<sup>1</sup> |  |
| --- | --- | --- | --- | --- | --- |
| Albumin [g/L] | 33,16 ± 0,97 | 27,96 ± 1,76 | 32,49 ± 1,69 | | - |
| Total protein [g/L] | 51,36 ± 1,63 | 50,83 ± 2,8 | 57,86 ± 1,88 | | - |
| Adiponectin [μg/mL] | 5,31 ± 0,44 | 6,4 ± 0,78 | 6,77 ± 0,79 | | - |
| AAT [mg/mL] | 1,85 ± 0,11 | 3,41 ± 0,31 | 3,03 ± 0,19 | | **B > A \*\*\***<br>**C > A \*\*\*** |
| A2Macro [mg/mL] | 2,5 ± 0,14 | 2,77 ± 0,28 | 2,11 ± 0,1 | | **B > C \*** |
| Lp(a) [μg/mL] | 257,55 ± 68,59 | 295,7 ± 66,69 | 153,74 ± 38,47 | | - |
| B2M [μg/mL] | 2,91 ± 0,29 | 9,57 ± 1,4 | 5,45 ± 0,86 | | **B > A \*\*\***<br>**B > C \*\*** |
| BDNF [ng/mL] | 11,78 ± 0,89 | 4,76 ± 0,6 | 10,41 ± 1,29 | | **A > B \*\*\***<br>**C > B \*\*** |
| CRP [μg/mL] | 43,1 ± 13,47 | 242,22 ± 37,04 | 189,81 ±<br>27,09 | | **B > A \*\*\***<br>**C > A \*\*** |
| C3 [mg/mL] | 1,21 ± 0,08 | 1,38 ± 0,12 | 1,36 ± 0,07 | | - |
| EN-RAGE<br>[ng/mL] | 250,77 ±<br>46,84 | 950,57 ± 153,36 | 871,77 ±<br>125,45 | | **B > A \*\***<br>**C > A \*\*** |
| Eotaxin-1<br>[pg/mL] | 154,58 ±<br>19,09 | 177,61 ± 32,18 | 169,2 ± 24,35 | | - |
| Factor VII<br>[ng/mL] | 290,96 ±<br>22,32 | 355,09 ± 32,06 | 338,37 ±<br>20,82 | | - |
| FRTN [ng/mL] | 1439,23 ±<br>428,85 | 2236,35 ±<br>484,33 | 2904,2 ±<br>641,14 | | - |
| Fibrinogen<br>[mg/mL] | 0,01 ± 0 | 0,04 ± 0,01 | 0,03 ± 0 | | **B > A \*\***<br>**C > A \*** |
| GM-CSF<br>[pg/mL] | 9,38 ± 0,38 | 9,52 ± 0,52 | 9,77 ± 0,77 | | - |
| Haptoglobin<br>[mg/mL] | 2,29 ± 0,35 | 5,46 ± 0,93 | 4,03 ± 0,46 | | **B > A \*\*** |
| IgA [mg/mL] | 3,33 ± 0,32 | 4,76 ± 0,62 | 3,31 ± 0,32 | | **B > C \*** |
| IgM [mg/mL] | 2,22 ± 0,2 | 3,46 ± 0,43 | 3,49 ± 0,33 | | **B > A \***<br>**C > A \*** |
| ICAM-1 [ng/mL] | 92,15 ± 9,43 | 199,61 ± 19,62 | 145,6 ± 22,86 | | **B > A \*\*** |
| IFN-gamma<br>[pg/mL] | 2,62 ± 0,12 | 2,5 ± 0 | 3,92 ± 0,96 | | - |
| IL-1 beta<br>[pg/mL] | 8,19 ± 0,7 | 11,01 ± 0,87 | 10,44 ± 0,56 | | **B > A \*** |

|  |  |  |  |  |
| --- | --- | --- | --- | --- |
| IL-1ra [pg/mL] | 490,46 ±<br>212,31 | 599,91 ± 172,38 | 600,11 ± 84,6 | - |
| IL-2 [pg/mL] | 26,6 ± 1,1 | 27,26 ± 1,76 | 25,5 ± 0 | - |
| IL-5 [pg/mL] | 5,83 ± 0,33 | 7,15 ± 1,32 | 5,5 ± 0 | - |
| IL-6 [pg/mL] | 22 ± 8,27 | 46,05 ± 13,28 | 108,16 ±<br>46,33 | - |
| IL-8 [pg/mL] | 42,79 ± 9,48 | 99,22 ± 18,82 | 63,1 ± 10,08 | **B > A \*** |
| IL-10 [pg/mL] | 25,19 ± 7,34 | 20,65 ± 3,22 | 20,23 ± 2,96 | - |
| IL-12p40<br>[ng/mL] | 0,59 ± 0,06 | 0,75 ± 0,06 | 0,69 ± 0,04 | - |
| IL-17 [pg/mL] | 1,58 ± 0,21 | 2 ± 0,31 | 1,81 ± 0,21 | - |
| IL-18 [pg/mL] | 332,48 ±<br>58,24 | 936,5 ± 187,53 | 855,4 ±<br>144,83 | **B > A \***<br>**C > A \*** |
| MIP-1 alpha<br>[pg/mL] | 26,81 ± 2,99 | 60 ± 9,45 | 29,09 ± 2,94 | **B > A \*\*\***<br>**B > C \*\*\*** |
| MIP-1 beta<br>[pg/mL] | 401,35 ±<br>56,26 | 686,78 ± 201,81 | 573,54 ±<br>63,52 | - |
| MMP-3 [ng/mL] | 7,37 ± 1,34 | 20,2 ± 3,43 | 8,66 ± 1,11 | **B > A \*\*\***<br>**B > C \*\*\*** |
| MMP-9 [ng/mL] | 35,5 ± 2,78 | 46,57 ± 3,49 | 38,74 ± 2,58 | **B > A \*** |
| MCP-1 [pg/mL] | 473,83 ±<br>60,27 | 957,74 ± 175,78 | 1274,91 ±<br>283,05 | **C > A \*** |
| Myoglobin<br>[ng/mL] | 108,69 ± 17,2 | 179,96 ± 15,58 | 109,31 ±<br>15,84 | **B > A \***<br>**B > C \*** |
| PAI-1 [ng/mL] | 217,85 ± | 253,41 ± 26,81 | 353,6 ± 33,72 | **C > A \*\*** |
|  | 22,86 |  |  |  |  |
| PARC [ng/mL] | 187,85 ±<br>31,35 | 319,52 ± 52,11 | 229,69 ±<br>24,21 |  | **B > A \*** |
| SAP [µg/mL] | 15,38 ± 0,93 | 24,17 ± 2,23 | 20,63 ± 1,02 |  | **B > A \*\*\***<br>**C > A \*** |
| SCF [pg/mL] | 438,98 ±<br>35,14 | 966,04 ± 98,35 | 563,67 ±<br>41,53 |  | **B > A \*\*\***<br>**B > C \*\*\*** |
| RANTES<br>[ng/mL] | 26,43 ± 3,01 | 16,88 ± 2,35 | 20,62 ± 1,71 |  | **A > B \*** |
| TBG [µg/mL] | 44,19 ± 1,83 | 48,52 ± 2,83 | 45,29 ± 2,34 |  | - |
| TIMP-1 [ng/mL] | 298,27 ±<br>38,77 | 474,87 ± 30,13 | 404,03 ±<br>39,84 |  | **B > A \*\*** |
| TNFR2 [ng/mL] | 110,88 ±<br>93,99 | 64,48 ± 8,43 | 34,87 ± 7,07 |  | - |
| VCAM-1<br>[ng/mL] | 947,04 ±<br>68,15 | 1568,09 ±<br>142,19 | 1291,77 ±<br>95,44 |  | **B > A \*\*\*** |
| VEGF [pg/mL] | 313,23 ±<br>44,04 | 421,7 ± 62,61 | 486,4 ± 49,45 |  | - |
| VDBP [µg/mL] | 250,31 ±<br>16,54 | 283,7 ± 26,06 | 281,37 ±<br>16,26 |  | - |
| vWF [µg/mL] | 343,92 ±<br>28,95 | 575,35 ± 45,67 | 440,06 ±<br>40,27 |  | **B > A \*\*** |
| AFP [ng/mL] | 14,28 ± 13,63 | 0,83 ± 0,16 | 0,56 ± 0,04 |  | - |
| ANG-1 [ng/mL] | 36,12 ± 2,83 | 32,79 ± 3,87 | 41,96 ± 3,39 |  | - |
| AXL [ng/mL] | 7,36 ± 0,62 | 10,66 ± 1,01 | 7,31 ± 0,73 |  | **B > A \*** |

|  |  |  |  |  |
| --- | --- | --- | --- | --- |
|  |  |  |  | **B > C \*** |
| CA-125 [U/mL] | 10,53 ± 3,8 | 35,70 ± 8,71 | 7,07 ± 1,26 | **B > A \*\***<br>**B > C \*\*\*** |
| CA-19-9 [U/mL] | 47,28 ± 20,17 | 33,14 ± 11,35 | 22,22 ± 5,97 | - |
| CA-9 [ng/mL] | 0,35 ± 0,09 | 0,53 ± 0,08 | 0,34 ± 0,05 | **B > C \*** |
| CEA [ng/mL] | 1,99 ± 0,95 | 3,84 ± 0,95 | 2,20 ± 0,33 | - |
| HCC-4 [ng/mL] | 4,26 ± 0,32 | 5,93 ± 0,39 | 3,04 ± 0,32 | **A > C \***<br>**B > A \*\***<br>**B > C \*\*\*** |
| Decorin [ng/mL] | 2,26 ± 0,19 | 3,21 ± 0,22 | 2,58 ± 0,22 | **B > A \*** |
| FAS [ng/mL] | 29,82 ± 3,84 | 66,26 ± 6,4 | 43,7 ± 17,63 | - |
| HGF [ng/mL] | 9,99 ± 0,94 | 36,75 ± 8,03 | 31,04 ± 8,28 | **B > A \*** |
| hCG [mIU/mL] | 3,19 ± 0,89 | 2,9 ± 0,37 | 3,63 ± 0,78 | - |
| IL-18bp [ng/mL] | 11,92 ± 0,89 | 23,61 ± 2,18 | 20,19 ± 1,84 | **B > A \*\*\***<br>**C > A \*\*** |
| MMP-1 [ng/mL] | 13,96 ± 1,92 | 24,58 ± 4,51 | 25,55 ± 3,82 | - |
| MMP-7 [ng/mL] | 7,17 ± 1,55 | 11,81 ± 1,4 | 7,97 ± 1,04 | **B > C \*** |
| MMP-9, total<br>[ng/mL] | 869,77 ±<br>93,18 | 899,91 ± 87,2 | 1247,29 ±<br>107,05 | **C > A \***<br>**C > B \*** |
| NSE [ng/mL] | 0,59 ± 0,06 | 1,03 ± 0,12 | 2,11 ± 0,22 | **C > A \*\*\***<br>**C > B \*\*\*** |
| PECAM-1<br>[ng/mL] | 54,85 ± 2,84 | 92,43 ± 9,08 | 77,23 ± 5,94 | **B > A \*\*\***<br>**C > A \*** |
| SP-D [ng/mL] | 15,71 ± 2,16 | 73,97 ± 14,57 | 34,19 ± 4,73 | **B > A \*\*\***<br>**B > C \*\*\*** |
| TRAIL-R3<br>[ng/mL] | 6,82 ± 0,6 | 16,24 ± 2,39 | 8,61 ± 0,76 |  | **B > A \*\*\***<br>**B > C \*\*\*** |

**Figure 1:**
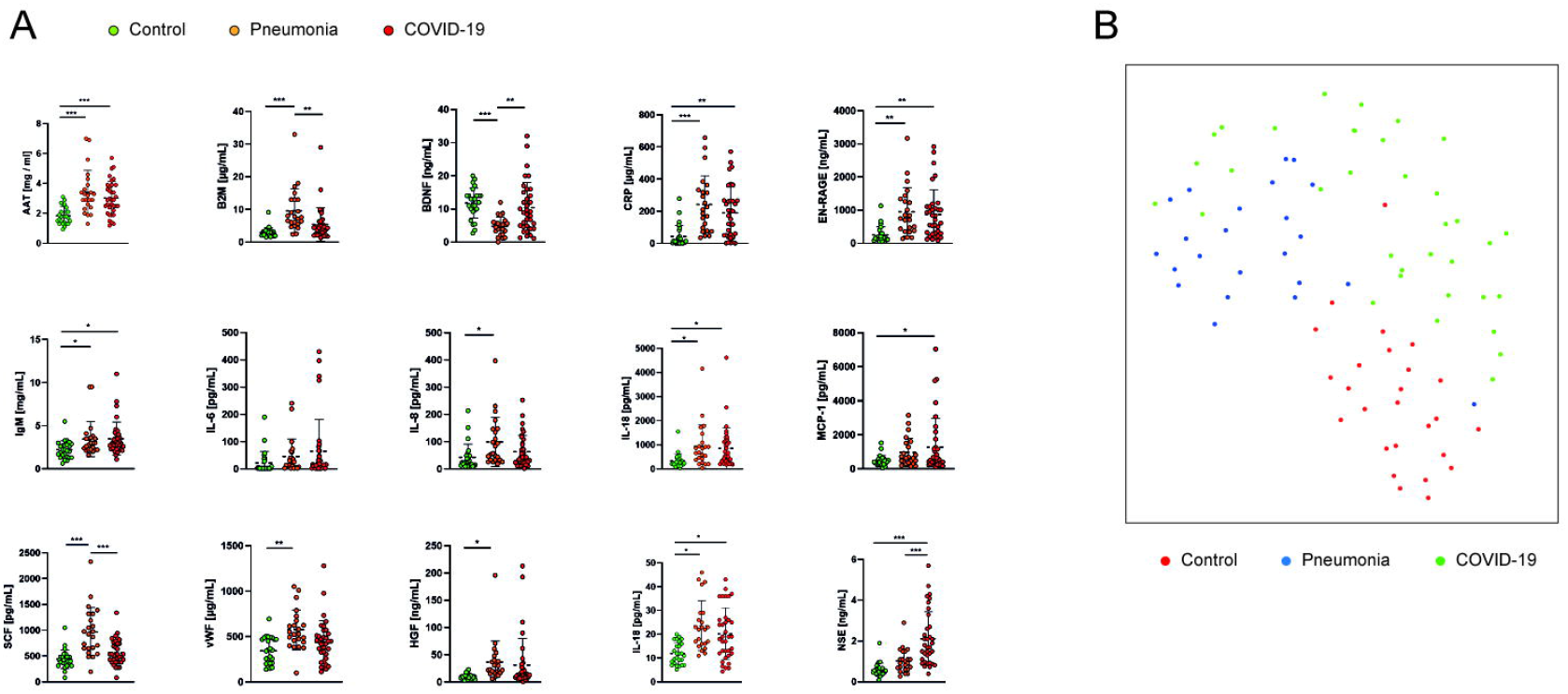
Serum markers discriminate between control, pneumonia, or COVID-19. (A) Serum concentrations of biomarkers that showed differences between control, pneumonia, or COVID-19 groups. p < 0.05, ** p < 0.005, *** p < 0.001 as calculated using one-way Analysis of Variance (ANOVA, assuming normality) with Bonferroni correction as post-hoc test. (B) UMAP dimension reduction integrating all detectable biomarkers shows segregation the three groups. We clustered the individuals by 3-means clustering and computed for each cluster how many individuals from the three groups were contained. Following this clustering we computed Fisher’s Exact test on the 3×3 contingency table, validating a significant clustering (p < 0.0001).

Comparing COVID-19 to conventional pneumonia revealed significant differences and concentrations of BDNF, MMP-9, and NSE were increased, while A2Macro, B2M, IgA, MIP1alpha, MMP-3, myoglobin, SCF, TNFR2, vWF, AXL, CA-125, CA-9, HCC-4, decorin, MMP-7, SPD, TRAIL-R3 were decreased in COVID-19 as compared to pneumonia patients. No significant differences were found for IL-6, or FRTN. As compared to the control group, COVID-19 patients showed increased concentrations of CRP, fibrinogen, IgM, PECAM-1, NSE, AAT, N-RAGE, IL-18, IL-18bp, MCP-1, PAI-1, and SAP. Using a UMAP approach, we classified individuals of all three groups based on their biomarker levels. This procedure resulted in significant segregation of the three groups (Fig. 1B).

### Factors associated with disease severity in COVID-19 patients

COVID-19 is a dynamic disease that often progresses towards respiratory failure and systemic inflammation and shock. The differences of blood concentrations between the disease severity stages (normal care vs. intensive care treatment) were analyzed (Table 3A). Figure 2A displays the significantly altered biomarkers. More severely sick patients requiring ICU treatment had increased levels of AAT, CRP, EN-RAGE, IL-1ra, IL-8, IL-10, IL-18, MCP-1, myoglobin, SAP, TIMP-1, vWF, decorin, and HGF, but decreased blood concentrations of albumin, total protein, adiponectin, A2Macro, BDNF, IL-17, and RANTES.

**Table 3:** Biomarkers significantly increased in patients with severe COVID-19 disease (A) or in patients who died during the course of disease. **(B)**. Data are median ± SD. P values were calculated using two-sided T-test.

| Analyte | Severe COVID-19 (A) |  | Survivor vs. Non-survivor (B) |  |
| --- | --- | --- | --- | --- |
|  | complicated | critical | Survivor | Non-Survivor |
| Albumin [g/L] | **35.13 ± 2.01** | **24.90 ± 2.06** | **35.13 ± 1.89** | **27 ± 3.14** |
|  | **p = 0.005** |  | **p = 0.026** |  |
| Total protein [g/L] | **61.65 ± 1.71** | **48.60 ± 3.80** | 60.39 ± 2.10 | 52.73 ± 3.72 |
|  | **p = 0.001** |  | p = 0.063 |  |
| Adiponectin [μg/mL] | **7.41 ± 0.93** | **4.13 ± 0.51** | **7.86 ± 1.12** | **4.77 ± 0.55** |
|  | **p = 0.032** |  | **p = 0.019** |  |
| AAT [mg/mL] | **2.7 ± 0.20** | **3.87 ± 0.32** | 3.01 ± 0.24 | 3.1 ± 0.34 |
|  | **p = 0.004** |  | p = 0.831 |  |
| A2Macro [mg/mL] | **2.24 ± 0.11** | **1.77 ± 0.24** | 2.20 ± 0.10 | 1.93 ± 0.24 |
|  | **p = 0.041** |  | p = 0.323 |  |
| Lp(a) [μg/mL] | 184.96 ± 56.54 | 99.40 ± 28.09 | 192.30 ± 56.36 | 80.55 ± 22.05 |
|  | p = 0.341 |  | p = 0.190 |  |
| B2M [μg/mL] | 5.52 ± 1.27 | 5.61 ± 0.84 | 4.79 ± 1.17 | 7.11 ± 1.17 |
|  | p = 0.964 |  | **p = 0.226** |  |
| BDNF [ng/mL] | **12.48 ± 1.60** | **4.22 ± 0.75** | **12.35 ± 1.73** | **6.02 ± 1.19** |
|  | **p = 0.00006** |  | **p = 0.005** |  |
| CRP [μg/mL] | **132.5 ± 28.92** | **333.30 ± 37.68** | 178.10 ± 33.76 | 226.09 ± 48.77 |
|  | **p = 0.0004** |  | p = 0.425 |  |
| C3 [mg/mL] | 1.36 ± 0.07 | 1.27 ± 0.14 | 1.39.00 ± 0.08 | 1.27 ± 0.12 |
|  | p = 0.515 |  | p = 0.403 |  |
| EN-RAGE [ng/mL] | **606.30 ± 123.08** | **1484.00 ± 250.40** | 812.00 ± 144.24 | 1048.73 ± 260.16 |
|  | **p = 0.001** |  | p = 0.395 |  |
| Eotaxin-1<br>[pg/mL] | 160.09 ± 30.63 | 198.20 ± 48.95 | **122.04 ± 17.62** | **269 ± 59.35** |
|  | p = 0.505 |  | **p = 0.036** |  |
| Factor VII<br>[ng/mL] | 348.43 ± 27.71 | 304.40 ± 34.48 | 320.91 ± 27.97 | 365.82 ± 29.33 |
|  | p = 0.365 |  | p = 0.330 |  |
| FRTN [ng/mL] | 2014.57 ±<br>709.46 | 4825.30 ±<br>1332.19 | 2037.35 ±<br>498.16 | 4964.09 ±<br>1629.9 |
|  | p = 0.051 |  | p = 0.112 |  |
| Fibrinogen<br>[mg/mL] | 0.04 ± 0.005 | 0.03 ± 0.003 | 0.03 ± 0.01 | 0.03 ± 0.00 |
|  | p = 0.325 |  | p = 0.198 |  |
| GM-CSF<br>[pg/mL] | 10.17 ± 1.17 | 9.00 ± 0.00 | 10.17 ± 1.17 | 9.00 ± 0.00 |
|  | p = 0.518 |  | p = 0.498 |  |
| Haptoglobin<br>[mg/mL] | 3.74 ± 0.60 | 4.32 ± 0.7 | 4.09 ± 0.65 | 4.26 ± 0.50 |
|  | p = 0.575 |  | p = 0.860 |  |
| IgA [mg/mL] | 3.00 ± 0.27 | 4.21 ± 0.87 | 3.40 ± 0.37 | 2.96 ± 0.64 |
|  | p = 0.210 |  | p = 0.535 |  |
| IgM [mg/mL] | 3.60 ± 0.41 | 3.45 ± 0.65 | 3.85 ± 0.45 | 2.68 ± 0.38 |
|  | p = 0.849 |  | p = 0.104 |  |
| ICAM-1 [ng/mL] | 108.70 ± 13.61 | 239.70 ± 66.94 | 119.74 ± 18.81 | 206 ± 58.97 |
|  | p = 0.085 |  | p = 0.189 |  |
| IL-1 beta<br>[pg/mL] | 10.44 ± 0.73 | 10.61 ± 1.06 | **9.62 ± 0.67** | **12.03 ± 0.98** |
|  | p = 0.899 |  | **p = 0.049** |  |
| IL-1RA [pg/mL] | **418.48 ± 74.94** | **1062.20 ±**<br>**172.41** | **441.39 ± 85.33** | **969.09 ± 153.41** |
|  | **p = 0.005** |  | **p = 0.003** |  |
| IL-6 [pg/mL] | 71.95 ± 56.84 | 211.40 ± 92.09 | 39.94 ± 17.32 | 260.38 ± 135.85 |
|  | p = 0.195 |  | p = 0.138 |  |
| IL-8 [pg/mL] | **35.76 ± 6.71** | **122.60 ± 21.63** | **39.28 ± 6.36** | **116.82 ± 22.12** |
|  | **p = 0.003** |  | **p = 0.006** |  |
| IL-10 [pg/mL] | **16.57 ± 3.67** | **30.60 ± 4.72** | **14.57 ± 2.72** | **33.45 ± 5.89** |
|  | **p = 0.036** |  | **p = 0.011** |  |
| IL-12p40<br>[ng/mL] | 0.73 ± 0.05 | 0.60 ± 0.10 | 0.65 ± 0.05 | 0.77 ± 0.09 |
|  | p = 0.188 |  | p = 0.180 |  |
| IL-17 [pg/mL] | **1.98 ± 0.30** | **1.30 ± 0.00** | 1.84 ± 0.28 | 1.60 ± 0.30 |
|  | **p = 0.033** |  | p = 0.600 |  |
| IL-18 [pg/mL] | **556.17 ± 83.49** | **1505.90 ± 395.08** | 634.09 ± 96.95 | 1373.00 ± 378.61 |
|  | **p = 0.041** |  | p = 0.085 |  |
| MIP-1 alpha<br>[pg/mL] | 27.13 ± 3.04 | 32.1 ± 7.18 | 26.04 ± 2.98 | 36.45 ± 6.66 |
|  | p = 0.535 |  | p = 0.175 |  |
| MIP-1 beta<br>[pg/mL] | 502.39 ± 71.82 | 690.5 ± 130.62 | 547.96 ± 74.92 | 660.36 ± 125.37 |
|  | p = 0.184 |  | p = 0.423 |  |
| MMP-3 [ng/mL] | 9.28 ± 1.59 | 7.95 ± 1.32 | 8.07 ± 1.19 | 10.17 ± 2.53 |
|  | p = 0.610 |  | p = 0.396 |  |
| MMP-9 [ng/mL] | 39.48 ± 3.40 | 36.1 ± 4.67 | 36.83 ± 3.39 | 44.82 ± 3.20 |
|  | p = 0.578 |  | p = 0.149 |  |
| MCP-1 [pg/mL] | **732.96 ± 292.33** | **2694.90 ± 512.90** | **576.35 ± 114.48** | **2836.09 ± 670.49** |
|  | **p = 0.001** |  | **p = 0.007** |  |
| Myoglobin<br>[ng/mL] | **77.91 ± 15.42** | **195.4 ± 28.03** | 93.35 ± 18.98 | 150.91 ± 27.54 |
|  | **p = 0.0004** |  | p = 0.094 |  |
| PAI-1 [ng/mL] | 305.83 ± 29.42 | 417.4 ± 77.93 | 354.39 ± 45.47 | 368.73 ± 49.57 |
|  | p = 0.108 |  | p = 0.848 |  |
| PARC [ng/mL] | 226.39 ± 32.54 | 253.20 ± 38.83 | 225.78 ± 29.96 | 228.73 ± 46.01 |
|  | p = 0.634 |  | p = 0.957 |  |
| SAP [µg/mL] | **18.43 ± 1.19** | **25.00 ± 1.12** | 20.13 ± 1.40 | 21.82 ± 1.44 |
|  | **p = 0.002** |  | p = 0.462 |  |
| SCF [pg/mL] | 556.43 ± 43.23 | 585.95 ± 108.10 | **473.15 ± 38.62** | **760.18 ± 78.54** |
|  | p = 0.761 |  | **p = 0.001** |  |
| RANTES<br>[ng/mL] | **22.78 ± 2.02** | **15.27 ± 3.03** | 20.05 ± 2.15 | 20.86 ± 3.04 |
|  | **p = 0.048** |  | p = 0.829 |  |
| TBG [µg/mL] | 46.17 ± 2.99 | 42.9 ± 4.39 | 44.78 ± 3.10 | 45.45 ± 3.71 |
|  | p = 0.547 |  | p = 0.898 |  |
| TIMP-1 [ng/mL] | **324.04 ± 33.80** | **598.4 ± 87.38** | **354.3 ± 37.08** | **526.82 ± 91.84** |
|  | **p = 0.001** |  | **p = 0.045** |  |
| TNFR2 [ng/mL] | 25.77 ± 5.56 | 55.2 ± 19.99 | 28.15 ± 5.86 | 51.27 ± 18.45 |
|  | p = 0.066 |  | p = 0.137 |  |
| VCAM-1<br>[ng/mL] | 1198.26 ±<br>119.11 | 1496.7 ± 165.26 | 1191.65 ±<br>117.04 | 1523.09 ±<br>166.03 |
|  | p = 0.168 |  | p = 0.115 |  |
| VEGF [pg/mL] | 456.65 ± 65.61 | 576.1 ± 81.39 | 450.26 ± 63.22 | 581.55 ± 80.43 |
|  | p = 0.300 |  | p = 0.229 |  |
| VDBP [µg/mL] | 278.13 ± 21.49 | 288.90 ± 29.08 | 273.26 ± 23.78 | 294.27 ± 14.49 |
|  | p = 0.778 |  | p = 0.456 |  |
| vWF [µg/mL] | **361.39 ± 33.61** | **632.5 ± 89.67** | 429.61 ± 54.04 | 481.36 ± 59.19 |
|  | **p = 0.001** |  | p = 0.563 |  |
| AFP [ng/mL] | 0.60 ± 0.06 | 0.49 ± 0.00 | 0.52 ± 0.03 | 0.54 ± 0.05 |
|  | p = 0.105 |  | p = 0.780 |  |
| ANG-1 [ng/mL] | 41.26 ± 2.72 | 34.55 ± 6.37 | **47.02 ± 4.51** | **32.45 ± 3.88** |
|  | p = 0.260 |  | **p = 0.048** |  |
| AXL [ng/mL] | 6.99 ± 0.67 | 7.75 ± 2.06 | 6.63 ± 0.68 | 8.84 ± 1.82 |
|  | p = 0.651 |  | p = 0.174 |  |
| CA-125 [U/mL] | 8.02 ± 1.83 | 5.67 ± 1.22 | 7.69 ± 1.86 | 5.69 ± 1.07 |
|  | p = 0.425 |  | p = 0.480 |  |
| CA-19-9 [U/mL] | 23.17 ± 8.6 | 24.14 ± 6.81 | 22.4 ± 8.54 | 23.54 ± 6.85 |
|  | p = 0.945 |  | p = 0.932 |  |
| CA-9 [ng/mL] | 0.34 ± 0.07 | 0.38 ± 0.09 | **0.26 ± 0.06** | **0.48 ± 0.08** |
|  | p = 0.747 |  | **p = 0.041** |  |
| CEA [ng/mL] | 1.92 ± 0.38 | 3.1 ± 0.65 | **1.58 ± 0.3** | **3.58 ± 0.68** |
|  | p = 0.112 |  | p = 0.017 |  |
| HCC-4 [ng/mL] | 3.05 ± 0.38 | 3.03 ± 0.71 | 2.69 ± 0.34 | 3.56 ± 0.69 |
|  | p = 0.976 |  | p = 0.211 |  |
| Decorin [ng/mL] | **2.28 ± 0.24** | **3.42 ± 0.42** | 2.27 ± 0.18 | 3.26 ± 0.53 |
|  | p = 0.018 |  | p = 0.102 |  |
| FAS [ng/mL] | 25.49 ± 4.08 | 91.43 ± 60.48 | 21.76 ± 3.14 | 91.64 ± 54.6 |
|  | p = 0.305 |  | p = 0.230 |  |
| HGF [ng/mL] | 15.88 ± 3.91 | 70.64 ± 23.99 | 16.47 ± 3.05 | 63.14 ± 23.43 |
|  | p = 0.049 |  | p = 0.076 |  |
| hCG [mIU/mL] | 3.74 ± 1.13 | 3.67 ± 0.86 | 3.87 ± 1.13 | 3.26 ± 0.78 |
|  | p = 0.967 |  | p = 0.727 |  |
| IL-18bp [ng/mL] | 18.18 ± 2.38 | 25.60 ± 2.67 | **17.52 ± 2.13** | **26.91 ± 2.89** |
|  | p = 0.075 |  | p = 0.016 |  |
| MMP-1 [ng/mL] | 24.98 ± 4.46 | 21.68 ± 4.7 | 24.27 ± 5.11 | 24.99 ± 5.09 |
|  | p = 0.661 |  | p = 0.931 |  |
| MMP-7 [ng/mL] | 6.47 ± 0.7 | 12.21 ± 2.91 | 6.85 ± 1.10 | 10.51 ± 2.26 |
|  | p = 0.084 |  | p = 0.109 |  |
| MMP-9, total<br>[ng/mL] | 1152.48 ±<br>126.91 | 1389.8 ± 225.57 | 1372.09 ±<br>130.57 | 1080.09 ±<br>172.68 |
|  | p = 0.336 |  | p = 0.201 |  |
| NSE [ng/mL] | 1.85 ± 0.27 | 2.79 ± 0.41 | 1.99 ± 0.28 | 2.37 ± 0.40 |
|  | p = 0.064 |  | p = 0.452 |  |
| PECAM-1<br>[ng/mL] | 69.52 ± 5.63 | 96.7 ± 15.09 | 74.04 ± 5.51 | 87.27 ± 14.74 |
|  | p = 0.118 |  | p = 0.416 |  |
| SP-D [ng/mL] | 31.47 ± 5.76 | 42.4 ± 9.47 | 34.64 ± 5.29 | 35.67 ± 10.33 |
|  | p = 0.317 |  | p = 0.922 |  |
| TRAIL-R3<br>[ng/mL] | 8.34 ± 0.92 | 9.94 ± 1.55 | 8.17 ± 0.88 | 9.04 ± 1.54 |
|  | p = 0.361 |  | p = 0.603 |  |

**Figure 2.**
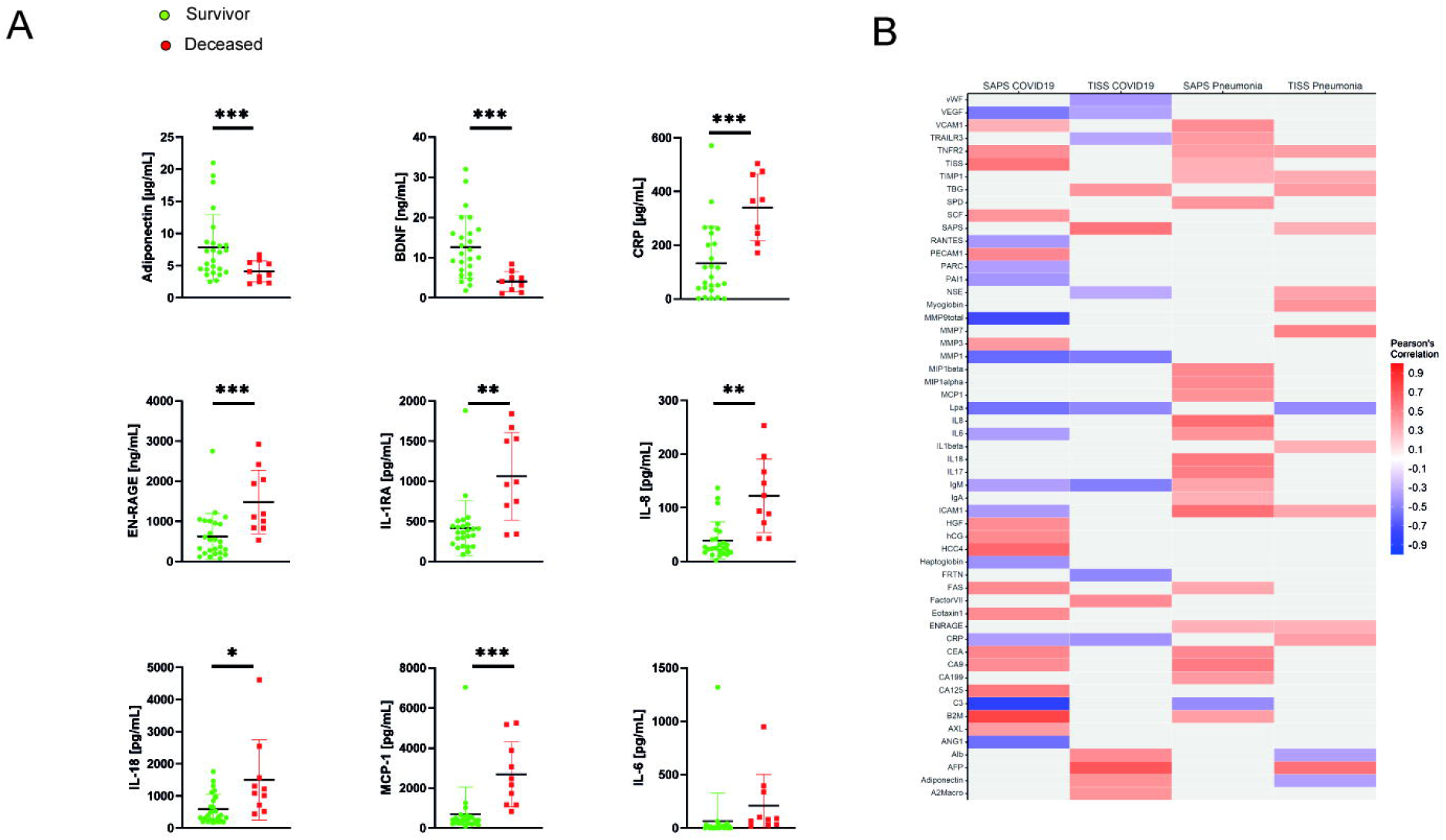
Biomarkers levels are associated with disease severity. (A) Biomarkers with significantly altered serum concentration in severe patients. p < 0.05, ** p < 0.005, *** p < 0.001 as calculated by two-sided T-test. (B) Biomarkers are associated with SAPS and TISS measurements; the magnitude of Pearson’s R is indicated by the color scheme.

To further evaluate the association between serum markers and disease severity, we measured the ICU scores SAPS II (Simplified Acute Physiology Score) and TISS (Therapeutic intervention scoring system) in COVID-19 patients. Table S2 presents results of bivariate correlation analysis, Figure 2B displays a summary correlation plot.

### Factors associated with death

One important goal of the use of biomarkers is to identify patients with increased risk of dying. As no death occurred in the control group and few in the pneumonia group, we identified factors associated with mortality only in the COVID-19 group (Table 3B).

Figure 3A displays the significantly changed biomarkers between the survivors and the deceased patients. T-test identified increased levels of eotaxin, FRTN, IL-1beta, IL-1ra, IL-8, IL-10, MCP-1, SCF, TIMP-1, CA-9, CEA, IL-18p. Decreased concentrations were found for albumin, adiponectin, BDNF, and ANG-1. No significant differences were found for BMI or age. A UMAP analysis (Fig. 3B) shows a significant separation between these two groups. A linear discriminant analysis was performed to test whether the serum markers identified above can be used to build a model that predicts mortality. The canonical discriminant functions revealed an Eigenvalue of 2.964 and a canonical correlation of 0.865 (Wilks-Lambda 0.252, Chi-quadrat P = 0.014) indicated a valid model with high standardized canonical discriminant function coefficients for IL-6 (0.532), IL-8 (0.467), MCP-1 (0.625), SCF (0.567), TIMP-1 (−0.920), decorin (−0.737), and HGF (0.614). This model resulted in a correct classification of 97.1 % of the original cases and 73.5 % of the cross-validation cases.

**Figure 3.**
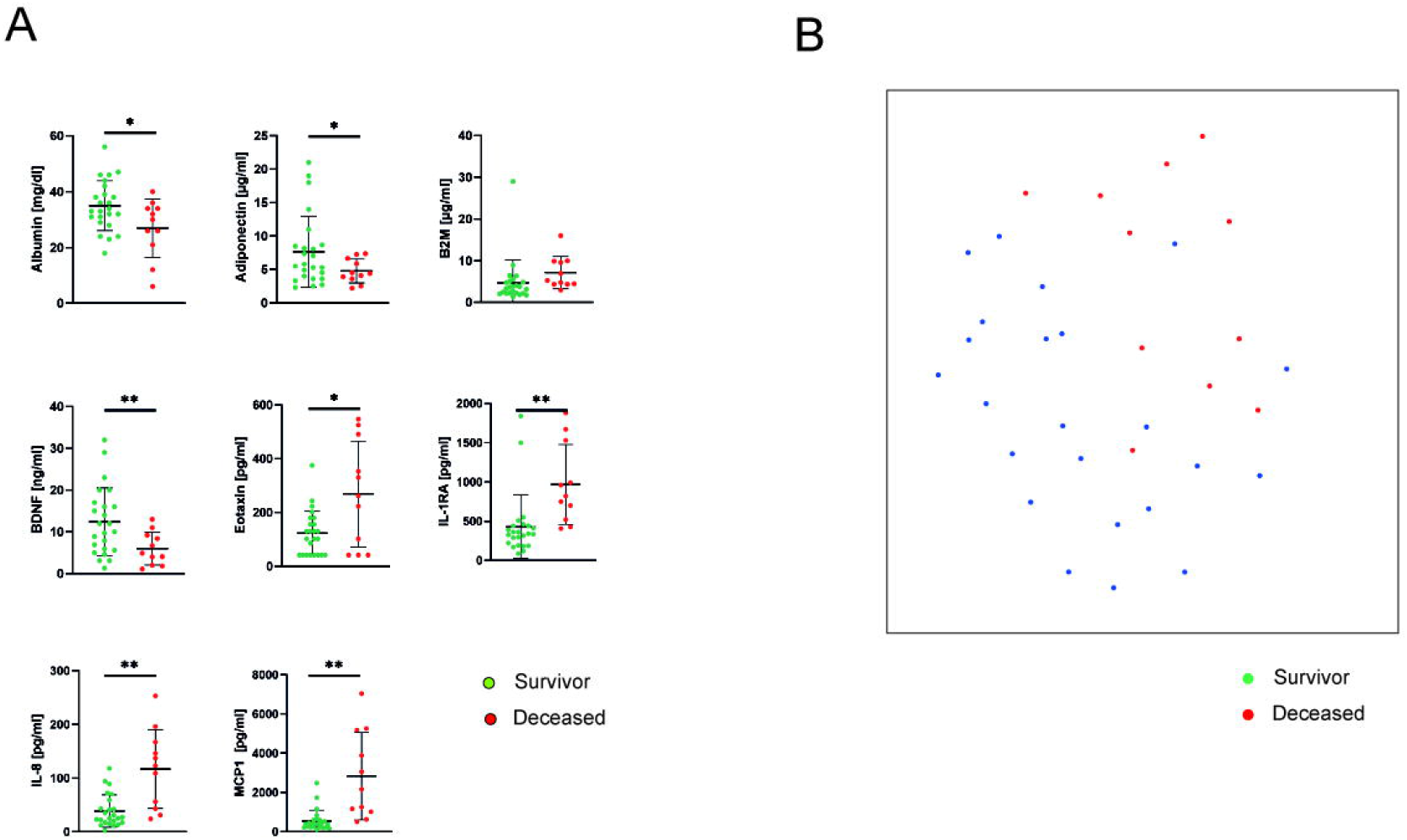
Biomarkers patterns are associated with mortality risk. (A) Serum markers with significantly altered serum concentration in patients who died during the course of disease. p < 0.05, ** p < 0.005, *** p < 0.001 as calculated by two-sided T-test. (B) UMAP analysis showed a significant separation of these two groups. We clustered the individuals by 2-means clustering and computed for each cluster how many individuals from the survival and non-survival group were contained in each cluster. Following this clustering we computed Fisher’s Exact test on the 2×2 contingency table, validating a significant clustering (p < 0.01).

While the number of cases is low, we used several rounds neural network analysis (SPPS module multilayer perceptron, MLP, one hidden layer) using “death” as dependent variable and the biomarkers with different serum concentrations in survivors vs. non-survivors as covariates. The analysis was performed four times using different random training sets and resulted in: 1. Analysis: correct prediction in the training / test set of 88.5 / 87.5 % (with the 5 most relatively important variables: IL-6, Eotaxin, IL-8, IL-1RA, MCP-); 2. Analysis 100 / 100 % correct prediction (MCP-1, SCF, BDNF, Eotaxin, Decorin); 3. Analysis 95.5 / 91.7 % correct prediction (BDNF, IL-10, CA9, MCP-1, Adiponectin); 4. Analysis 95.8 / 100 % correct prediction (SCF, BDNF, IL18bp, IL-8, Age).

Finally, we performed a ROC analysis and found that IL-1ra, IL-8, IL-10, MCP-1, SCF and CA-9 have an AUC > 0.8. When these variables are subdivided in tertials and a score is established from these data, the AUC is 0.929.

## Discussion

The main findings of this study are that severe COVID-19 differs from conventional pneumonia by a specific blood cytokine profile. Several serum mediators are correlated with disease severity and outcome. While some blood biomarkers are significantly elevated in severe COVID-19, a classical “cytokine storm” as seen in other conditions was not observed. Several biomarkers are associated with disease severity and risk of death.

Blood biomarkers have been used to determine disease severity or risk for deleterious outcome in conventional pneumonia [11–13]. Biomarkers such as CRP, procalcitonin, and others are important tools for decision making in pneumonia patients. The identification of diagnostic, predictive or prognostic biomarkers is even more relevant in COVID-19.

There are two approaches to study the inflammatory processes and to identify biomarkers in COVID-19. One is the application of OMICs technologies to array multiple layers of gene expression, protein and metabolome distribution. A multi-OMICs approach identified patterns that separate mild from severe disease [9]. The combination of transcriptomics, proteomics and metabolomics revealed 219 molecular features related to COVID-19 and linked to complement activation, dysregulated lipid metabolism, and neutrophil activation [4]. A dysregulation of the myeloid cell compartment was found by transcriptomics and single-cell-analysis [2].

The other approach focuses on the measurement of selected biomarkers in clinically relevant cohorts. This approach does not allow a deeper insight into mechanisms but aids to quickly develop clinically relevant information. IL-1RA, IL-10, and RANTES were found to associate with disease severity in a longitudinal investigation [14]. IFN-γ and type I IFN production were found to be relatively deceased, while TNF-α, IL-6, and IL-8 were increased for a prolonged time [15]. The serial analysis of several cytokines in 113 patients showed the early rise in markers to be associated with worse outcome and an activation of type 2 cytokines in severe COVID-19 [3]. Measurement of IL-6, IL-8, TNF-α, and IL-1β were found to be predictive for severity and survival [8]. The measurement of a selected cytokine panel showed that COVID-19 is distinct from conventional pneumonia [7].

The present work applied a large panel of biomarkers to COVID-19 patients and control groups. The marker panel applied in the present study consists of mediators of inflammation and immunity, most of which are not yet part of clinical routine testing but have been shown to partake in cellular or humoral immune responses. The present study confirmed several of these findings and in addition shows a more comprehensive illustration of COVID-19 systemic inflammation. The blood marker profile of COVID-19 patients differs significantly from control patients with elective non-pulmonary surgery or severe pneumonia. The data show that COVID-19 is associated with a broad and complex activation of innate and adaptive immune responses.

In addition to several studies [10, 16], the present data showed increased serum concentrations of NSE, AAT, B2M, EN-RAGE, IL-18, MCP-1, PAI-1, SAP, IL-18bp, and MMP-9. Several newly identified blood markers were increased in patients with severe disease (AAT, EN-RAGE, ICAM-1, myoglobin, SAP, TIMP-1, vWF, decorin, HGF, MMP7, PECAM-1) or in patients that died during the disease course (FRTN, SCF, TIMP-1, CA-9, CEA, decorin, HGF).

The term “cytokine storm” in general describes the finding of an exaggerated immune response with excessively elevated cytokine levels and deleterious side effects [6] and it was often highlighted that this phenomenon would be a hallmark of COVID-19. Other reports questioned the role of a cytokine storm in COVID-19 in comparison to other inflammatory conditions [5]. Also, the preset results support this view with COVID-19 patients having a distinct pattern of increased inflammatory mediators but not an out-of-proportion increase.

The present study has limitations and strengths: The use of established, rapid multiplex assays allows to translate these findings to validation studies and clinical practice. One important aspect is the direct comparison to non-COVID-19 pneumonia and a control group. The present study has included relatively few COVID-patients and does not cover very mild or asymptomatic individuals nor sequential sampling. Overfitting might be a result of low patient numbers and validation in other cohorts is mandatory.

In conclusion, COVID-19 shows a distinct inflammatory blood marker profile separate from conventional pneumonia and extrapulmonary disease. Several markers were associated with disease severity and lethal outcome. We recommend a thorough investigation which of these biomarkers might be causative for immune dysregulation in severe COVID-19 or just a by-product of immune activation. Ideally, proteins strongly connected to COVID-19 pathophysiology could reveal potential targets for immunomodulating therapies for patients at risk.

## Data Availability

All data concerning this manuscript is available within the manuscript or its supplement.

## Acknowledgments

This work was supported by grants of the Rolf M. Schwiete Stiftung (2020-013), the Saarland University, and The State of Saarland. Protein biomarker assays have been performed in collaboration with Myriad RBM.

