## Supplemental Tables S1-S2 for "Distinct patterns of blood cytokines beyond a cytokine storm predict mortality in COVID-19"

**Table S1:** Clinical characteristics of all patients

| Nr | Diagnosis group (Control, pneumonia, COVID-19) | Diagnosis | Age  (rounded to 5 years interval) | Outcome  1=survivor, 2=Deceased |
| --- | --- | --- | --- | --- |
| 1 | Control | Cholangiocarcinoma | 75 | 1 |
| 2 | Control | Pancreatic cancer | 70 | 1 |
| 3 | Control | Hepatocelullar carcinoma | 70 | 1 |
| 4 | Control | Crohn’s disease, small intestinal resection | 55 | 1 |
| 5 | Control | Infection with uncertain focus | 45 | 1 |
| 6 | Control | Infrarenal aortic aneurysm | 65 | 1 |
| 7 | Control | Pancreatic cancer | 65 | 1 |
| 8 | Control | Colorectal carcinoma | 50 | 1 |
| 9 | Control | Hepatocelullar carcinoma | 70 | 1 |
| 10 | Control | Klatskin cancer | 55 | 1 |
| 11 | Control | Liver resection, liver cirrhosis | 55 | 1 |
| 12 | Control | Gastric carcinoma | 50 | 1 |
| 13 | Control | Non-Hodgkin lymphoma | 40 | 1 |
| 14 | Control | Colorectal carcinoma | 60 | 1 |
| 15 | Control | Gastric cancer | 90 | 1 |
| 16 | Control | Gastric bypass | 50 | 1 |
| 17 | Control | Prostate cancer | 55 | 1 |
| 18 | Control | Cholangiocarcinoma | 70 | 1 |
| 19 | Control | Reconnection operation after Hartmann-Operation | 60 | 1 |
| 20 | Control | Cholecystectomy | 45 | 1 |
| 21 | Control | Colorectal carcinoma | 70 | 1 |
| 22 | Control | Hepatocelullar carcinoma | 80 | 1 |
| 23 | Control | Perforated gastric ulcer | 70 | 1 |
| 24 | Control | Colon carcinoma with hepatic metastasis | 60 | 1 |
| 25 | Control | Colon carcinoma | 60 | 1 |
| 26 | Control | Whipple`s operation, papillary carcinoma | 70 | 1 |
| 27 | Pneumonia | - | 45 | 1 |
| 28 | Pneumonia | - | 55 | 1 |
| 29 | Pneumonia | - | 70 | 1 |
| 30 | Pneumonia | Influenza | 70 | 1 |
| 31 | Pneumonia |  | 70 | 1 |
| 32 | Pneumonia | Influenza | 70 | 1 |
| 33 | Pneumonia | - | 65 | 1 |
| 34 | Pneumonia | Aspiration pneumonia | 95 | 2 |
| 35 | Pneumonia | - | 80 | 2 |
| 36 | Pneumonia | - | 75 | 1 |
| 37 | Pneumonia | - | 70 | 1 |
| 38 | Pneumonia | - | 65 | 1 |
| 39 | Pneumonia | - | 75 | 1 |
| 40 | Pneumonia | - | 70 | 2 |
| 41 | Pneumonia | - | 85 | 1 |
| 42 | Pneumonia | AIDS | 55 | 1 |
| 43 | Pneumonia | - | 60 | 1 |
| 44 | Pneumonia | - | 40 | 1 |
| 45 | Pneumonia | - | 70 | 1 |
| 46 | Pneumonia | - | 35 | 1 |
| 47 | Pneumonia | - | 65 | 1 |
| 48 | Pneumonia | - | 60 | 1 |
| 49 | Pneumonia | - | 35 | 1 |
| 50 | COVID-19 | COVID-19 | 85 | 2 |
| 51 | COVID-19 | COVID-19 | 50 | 1 |
| 52 | COVID-19 | COVID-19 | 70 | 2 |
| 53 | COVID-19 | COVID-19 | 25 | 1 |
| 54 | COVID-19 | COVID-19 | 70 | 1 |
| 55 | COVID-19 | COVID-19 | 85 | 1 |
| 56 | COVID-19 | COVID-19 | 60 | 2 |
| 57 | COVID-19 | COVID-19 | 90 | 1 |
| 58 | COVID-19 | COVID-19 | 90 | 1 |
| 59 | COVID-19 | COVID-19 | 80 | 1 |
| 60 | COVID-19 | COVID-19 | 75 | 1 |
| 61 | COVID-19 | COVID-19 | 50 | 1 |
| 62 | COVID-19 | COVID-19 | 65 | 1 |
| 63 | COVID-19 | COVID-19 | 40 | 1 |
| 64 | COVID-19 | COVID-19 | 55 | 1 |
| 65 | COVID-19 | COVID-19 | 75 | 2 |
| 66 | COVID-19 | COVID-19 | 80 | 1 |
| 67 | COVID-19 | COVID-19 | 30 | 1 |
| 68 | COVID-19 | ARDS by COVID-19 | 60 | 2 |
| 69 | COVID-19 | COVID-19 | 60 | 1 |
| 70 | COVID-19 | COVID-19 | 80 | 2 |
| 71 | COVID-19 | COVID-19 | 80 | 1 |
| 72 | COVID-19 | COVID-19 | 70 | 1 |
| 73 | COVID-19 | COVID-19 | 90 | 1 |
| 74 | COVID-19 | COVID-19 | 15 | 1 |
| 75 | COVID-19 | ARDS by COVID-19 | 55 | 2 |
| 76 | COVID-19 | ARDS by COVID-19 | 55 | 1 |
| 77 | COVID-19 | ARDS by COVID-19 | 65 | 1 |
| 78 | COVID-19 | ARDS by COVID-19 | 65 | 1 |
| 79 | COVID-19 | ARDS by COVID-19 | 50 | 2 |
| 80 | COVID-19 | ARDS by COVID-19 | 85 | 2 |
| 81 | COVID-19 | ARDS by COVID-19 | 50 | 1 |
| 82 | COVID-19 | ARDS by COVID-19 | 70 | 2 |
| 83 | COVID-19 | ARDS by COVID-19 | 55 | 1 |
| 84 | COVID-19 | ARDS by COVID-19 | 60 | 2 |

**Table S2: Correlation of TISS and SAPS scores with biomarkers.** Upper line – Pearson’ R, lower line 2-sided significance

|  | Pneumonia | | COVID-19 | |
| --- | --- | --- | --- | --- |
| Analyte | SAPS | TISS | SAPS | TISS |
| IL-8 | ***0.592***  ***0.003*** | 0.246  0.258 | 0.131  0.684 | 0.056  0.864 |
| IL-17 | ***0.517***  ***0.012*** | 0.234  0.283 | -0.209  0.514 | -0.068  0.833 |
| IL-18 | ***0.541***  ***0.008*** | 0.254  0.249 | 0.119  0.514 | -0.068  0.833 |
| MIP-1a | ***0.472***  ***0.023*** | -0.154  0.484 | 0.282  0.374 | 0.283  0.373 |
| MIP-1b | ***0.481***  ***0.020*** | 0.102  0.642 | -0.022  0.946 | -0.094  0.771 |
| MCP-1 | ***0.437***  ***0.037*** | 0.01  0.963 | 0.02  0.95 | -0.145  0.653 |
| VCAM-1 | ***0.457***  ***0.028*** | 0.126  0.568 | 0.304  0.336 | -0.129  0.69 |
| CA-9 | ***0.534***  ***0.009*** | 0.076  0.73 | 0.456  0.136 | -0.006  0.985 |
| CRP | 0.162  0.46 | ***0.451***  ***0.031*** | 0.017  0.96 | 0.374  0.257 |
| LP(a) | 0.23  0.292 | ***-0.466***  ***0.025*** | -0.573  0.052 | -0.477  0.117 |
| Myoglobin | 0.127  0.564 | ***0.428***  ***0.042*** | 0.23  0.473 | -0.007  0.982 |
| AFP | 0.15  0.494 | ***0.571***  ***0.004*** | 0.133  0.68 | ***0.704***  ***0.011*** |
| ANG-1 | -0.074  0.739 | -0.172  0.433 | ***-0.587***  ***0.045*** | -0.218  0.495 |
| CEA | ***0.474***  ***0.022*** | 0.22  0.313 | 0.485  0.11 | -0.055  0.865 |
| Thromb | -0.369  0.084 | 0.027  0.903 | ***0.735***  ***0.01*** | ***-0.631***  ***0.038*** |
| Krea | 0.083  0.708 | -0.061  0.783 | ***0.685***  ***0.02*** | 0.059  0.862 |
| Urea | 0.214  0.327 | 0.114  0.605 | ***0.634***  ***0.036*** | 0.058  0.862 |
| B2M | 0.381  0.073 | -0.02  0.927 | ***0.779***  ***0.003*** | 0.29  0.361 |
| C3 | ***-0.460***  ***0.027*** | 0.044  0.84 | ***-0.857***  ***< 0.0001*** | -0.276  0.385 |
| HCC-4 | 0.003  0.989 | 0.258  0.235 | ***0.641***  ***0.034*** | 0.159  0.622 |
| MMP-1 | 0.3  0.164 | 0.023  0.917 | -***0.611***  ***0.035*** | -0.543  0.068 |
| MMP-7 | 0.168  0.143 | ***0.505***  ***0.014*** | 0.299  0.345 | 0.247  0.44 |
| MMP-9 | -0.279  0.197 | -0.165  0.451 | ***-0.740***  ***0.006*** | -0.166  0.607 |
| ICAM-1 | ***0.584***  ***0.003*** | 0.347  0.105 | -0.394  0.205 | -0.206  0.521 |
| SP-D | ***0.418***  ***0.047*** | 0.028  0.9 | 0.121  0.708 | 0.062  0.848 |
| Age | 0.035  0.888 | -0.308  0.153 | 0.407  0.189 | 0.268  0.4 |
